# Synthesis and new evidence from the PROTECT UK National Core Study: Determining occupational risks of SARS-CoV-2 infection and COVID-19 mortality

**DOI:** 10.1101/2023.06.30.23292079

**Authors:** S Rhodes, S Beale, M Cherrie, W Mueller, F Holland, M Matz, I Basinas, J Wilkinson, M Gittins, B Farrell, A Hayward, N Pearce, M van Tongeren

## Abstract

**Introduction:** The PROTECT National Core Study was funded by the UK Health and Safety Executive (HSE) to investigate routes of transmission for SARS-CoV-2 and variation between settings.

**Methods:** A workshop was organised in Oct 2022.We brought together evidence from five published epidemiological studies that compared risks of SARS-CoV-2 infection or COVID-19 mortality by occupation or sector funded by PROTECT relating to three non-overlapping data sets, plus additional unpublished analyses relating to the Omicron period. We extracted descriptive study level data and model results. We investigated risk across four pandemic waves using forest plots for key occupational groups by time-period.

**Results:** Results were largely consistent across different studies with different expected biases. Healthcare and social care sectors saw elevated risks of SARS-CoV-2 infection and COVID-19 mortality early in the pandemic, but thereafter this declined and varied by specific occupational subgroup. The education sector saw sustained elevated risks of infection after the initial lockdown period with little evidence of elevated mortality.

**Conclusions:** Increased in risk of infection and mortality were consistently observed for occupations in high risk sectors particularly during the early stage of the pandemic. The education sector showed a different pattern compared to the other high risk sectors, as relative risk of infections remained high in the later phased of the pandemic, although no increased in COVID-19 mortality (compared to low-risk occupations) was observed in this sector in any point during the pandemic.

## Background

SARS-CoV-2 infection was spreading in the UK from January 2020 with the first recorded death from COVID-19 in March of the same year. Understanding the role of the workplace in the risk of COVID-19 is important but complex. We cannot run controlled experiments that would provide the strongest evidence for causal relationships. We are reliant on observational data to understand how attending the workplace is associated with the risks from COVID-19, where bias is a concern (for example those attending a particular workplace may also be subject to particular risks/behaviours outside the workplace). In addition, the mitigations put in place in the UK to reduce COVID-19 transmission have varied over time, and between different parts of the UK. Many of these mitigations would be expected to affect workplace transmission and apply inconsistently to different sectors both in terms of uptake and affect – e.g., lockdown, vaccinations, mask use, use of lateral flow tests, homeworking. It seems likely that these interventions have contributed to between-occupational differences in risk. However, differences in risk may also be because of immunity from prior infections and periodic effects due to an undulating background infection rate; it is extremely difficult to specifically attribute changes over time to the mitigations in place.

HM Government funded the UK Health and Safety Executive (HSE) to lead a National Core Study called PROTECT (Partnership for Research in Occupational, Transport and Environmental COVID Transmission) from September 2020 in order to provide evidence-based guidance to protect the population from the virus. The overarching aim was to investigate how SARS-CoV-2 is transmitted from person to person, and how this varies in different settings and environments including the workplace.

PROTECT has funded a number of studies considering the effect of working in a particular occupation or sector during the pandemic. Each study has a different underlying methodology while attempting, as far as possible, to follow a common framework (1) . We know that bias is a concern in every observational study and that the direction of bias varies by underlying design and the data used. Consolidating evidence from different studies impacted by different biases can help to clarify between-occupational differences in COVID-19 outcomes and their risk factors. Combining evidence from multiple sources (2) has allowed us to strengthen conclusions and highlight uncertainties and areas for future research.

## Aims

In this piece of work we aimed to critique and synthesise existing analyses relating occupational risks of SARS-CoV-2 infection and COVID-19 mortality funded by the PROTECT National Core Study via the following research questions:

a. To what extent do a set of studies relating to differences between occupations and sectors in SARS-CoV-2 infection and COVID-19 mortality in the UK agree?
b. How do differences between occupations and sectors in SARS-CoV-2 infection and COVID-19 mortality change over time?
c. What are the expected biases in these studies, and how confident can we be in their overall results?

## Methods

In order to assess the key biases and strengths of the different studies and interpret our combined results we brought together study authors in an in-person workshop in October 2022 with seven study authors and a neutral scribe/facilitator. The format of the workshop included a presentation on study methodology followed by joint tabulation of key strength and biases, plus discussion and consensus on the key conclusions that could be made based on the combined evidence.

We brought together evidence from five published epidemiological studies that compared risks of SARS-CoV-2 infection and/or COVID-19 mortality by occupation or sector conducted with funding from PROTECT (ONS CIS, Virus Watch, ONS Mortality, ONS Excess Mortality, ONS Proportionate Mortality). Data extraction was carried out prior to the workshop by a researcher not involved in the original studies. Descriptive study level data on infection/mortality data source, age range, time period covered, region and covariate adjustment set was extracted from the published papers.

In order to allow comparison of outcome data from multiple studies we used a meta-analysis approach, but avoided pooling results due to expected heterogeneity in outcome definition, effect measures and design. We extracted relative effect sizes and 95% confidence intervals from studies comparing occupational risks to a reference category (e.g. non-essential workers) which we used to create forest plots for four key sectors previously thought to be at high risk: healthcare, social care, education and transport. We split results by COVID-19 wave using the following definitions: Wave 1 (January 2020 to November 2020) characterised by dominance of wild-type SARS-CoV-2 and stringent public health restrictions during periods of high community transmission; Wave 2 (December 2020 to May 2021), characterised by dominance of the alpha variant and stringent restrictions; Wave 3 (June 2021 to November 2021), characterised by dominance of the Delta variant and relaxation of public health restrictions); and Wave 4 (December 2021 onwards) characterised by dominance of the Omicron variant and further relaxation of remaining public health restrictions. Dates used to group results varied somewhat across studies and results were allocated to the wave with the greatest time overlap to the definition above. Waves 1 and 2 were amalgamated into a single period for infection risk analyses due to limited availability of Wave 1 data and inability to distinguish between these periods for serological results in some studies.

For two studies (3, 4) we ran new analyses including recent data to allow us to include the Omicron wave in our synthesis and details of these are in the appendix. A study comparing excess mortality by occupation (5, 6) did not report relative effect sizes or inferential statistics and therefore could not be included on forest plots – for this study we refer to the figures in the original publications and do not make direct comparisons to the other studies.

Table 1 provides a description of the five studies included in our analysis/synthesis. Two studies compared risks of SARS-CoV-2 infection(4, 7). The study relating to the Office of National Statistics Coronavirus Infection Survey (ONS CIS) uses data from a large national longitudinal cohort. This survey utilises random sampling designed to be representative of the UK population and data from survey specific repeated PCR tests to establish infection status. We include new results from this study in S1. The Virus Watch longitudinal cohort is smaller and used advertisements to recruit volunteers; infection status was determined using a combination of self-reported test results and linked NHS data, with serological testing amongst a sub-cohort. Three studies compared risks of COVID-19 mortality (3, 5, 8). One study linked ONS mortality data to UK census data from 2010 to allow analysis of the full UK population (aged 40 to 65). One study used the same ONS mortality data but used a proportionate mortality approach comparing deaths from COVID-19 to deaths from other causes. New analysis relating to this study are included in S2. Another study also used the same ONS mortality data but adopted an excess mortality approach, comparing deaths in 2020 and 2021 to deaths over a pre-pandemic period from 2015 to 2019 (6).

**Table 1:** Description of epidemiological studies relating occupational risks of SARS-CoV-2 infection and COVID-19 mortality funded by the PROTECT National Core Study

| Occupational risks of SARS-CoV-2 infection and COVID-19 mortality |  |  |  |  |  |  |  |
| --- | --- | --- | --- | --- | --- | --- | --- |
| Study short name | Reference | Data Source | Outcome effect measure | Age range | National region | Time periods# | Reference group# |
| ONS CIS | Rhodes 2022(18)* | ONS CIS | Infection hazard ratio | 20-64 | England, Scotland and Wales | W1 & W2: April 2020 to Feb 2021<br>W3: March 2021 to Dec 2021<br>W4: Jan 2022 to Aug 2022 | Non essential workers |
| Virus Watch | Beale 2022(7) | Virus Watch cohort | Infection risk ratios | 16 and over | England and Wales | W1 & W2: February 2020 to May 2021<br>W3 June 2021 to November 2021<br>W4 December 2021 to April 2022 | Other professional and associate occupations |
| ONS Mortality | Nafilyan 2021(8) | ONS mortality linked to 2010 census | Mortality hazard ratios | 40-64 | England and Wales | 24 January 2020 to 28 December 2020 | Non essential workers |
| ONS proportionate mortality | Cherrie 2022(3)* | ONS mortality | Proportionate mortality odds ratios | 20-64 | England and Wales | W1: Jan 2020 to Sept 2020<br>W2: Oct 2020 to May 2021<br>W3: June 2021 to October 2021<br>W4: Jan 2022 to June 2022* | Non essential workers |
| ONS excess mortality | Matz 2022(5) and Matz 2023(6) | ONS mortality | Excess mortality | 20-64 | England and Wales | 2020-2021 | Same group over previous 5 years |

\*Additional analyses included to incorporate data since Nov 2021.
#Time periods and reference groups utilised in figures. Source papers may include additional/alternative time periods and reference groups.

## Results

Figure 1 shows SARS-CoV-2 relative effects and confidence intervals relating to SARS-CoV-2 data infection and mortality data from for categories relating to health care. Both studies ONS CIS and Virus Watch saw elevated risks for healthcare workers during Wave 1 and 2 (combined) when compared to other occupations. During Wave 3 these elevated risks were reduced. Results from the ONS-CIS study suggested that risk of infection for healthcare associates and support workers were reduced compared to other workers. However, during Wave 4, the relative risk of infection for health care workers appeared to have slightly increased compared to non-essential or other workers, although the relative risks were small compared to wave 1 and 2.

**Fig 1:**
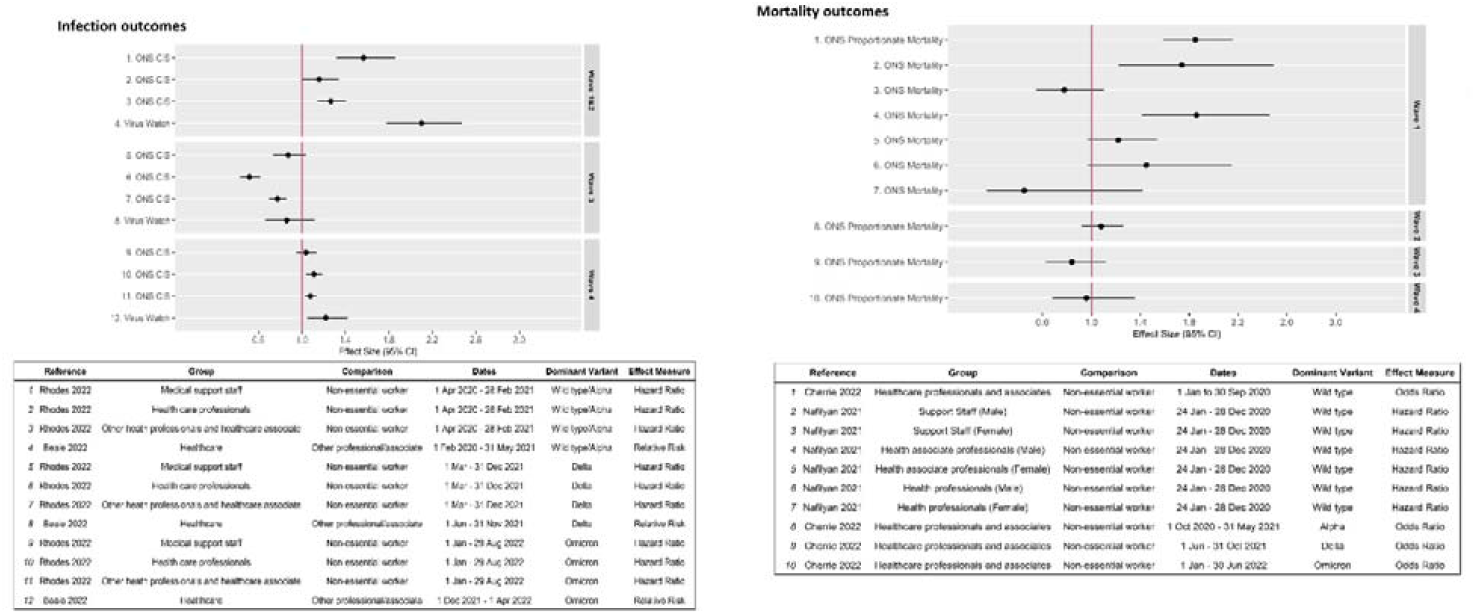
Effect size with 95% confidence interval for infection and mortality outcomes by COVID-19 wave for healthcare workers in the UK.

Relative effects and confidence intervals relating to SARS-CoV-2 mortality data from ONS mortality data based on two different analyses revealed patterns that were largely consistent with those seen for infection data. Elevated risks of mortality were seen during Wave 1 for most groups of healthcare worker with no evidence of increased mortality for healthcare workers in later waves. The ONS excess mortality, which covered 2020 and 2021, saw highest levels of excess mortality for healthcare workers that peaked in April 2020 followed by smaller peaks in excess mortality Aug 2020 to March 2021.

Figure S3 shows relative effects relating to SARS-CoV-2 data infection data from ONS CIS and Virus Watch for social care workers. Results from both studies are very consistent with elevated risks evident for combined Wave 1 and Wave 2 data, but no evidence of elevated risks during wave 3 or wave 4. Figure S4 suggests elevated mortality risks for social care workers in Waves 1, 2 and 4, with little evidence of increased mortality when compared to other workers in Wave 3. The excess mortality analysis suggested excess mortality for social care workers leaked later than for healthcare workers with the highest peak in March 2021.

Figure 2 shows relative effects and 95% confidence intervals for workers in the education sector. Results on infection relating to analysis of ONS CIS data and Virus Watch data were consistent and showed elevated risks during the combined wave 1 and 2 time period, Wave 3 and Wave 4, albeit with wide confidence intervals. We did not observe any evidence of increased mortality for workers in the education sector (relative to other workers) during any wave, with a slight reduction in the proportionate odds of mortality during wave 4. Excess mortality saw levels of excess mortality lower than those for other types of key worker.

**Fig 2:**
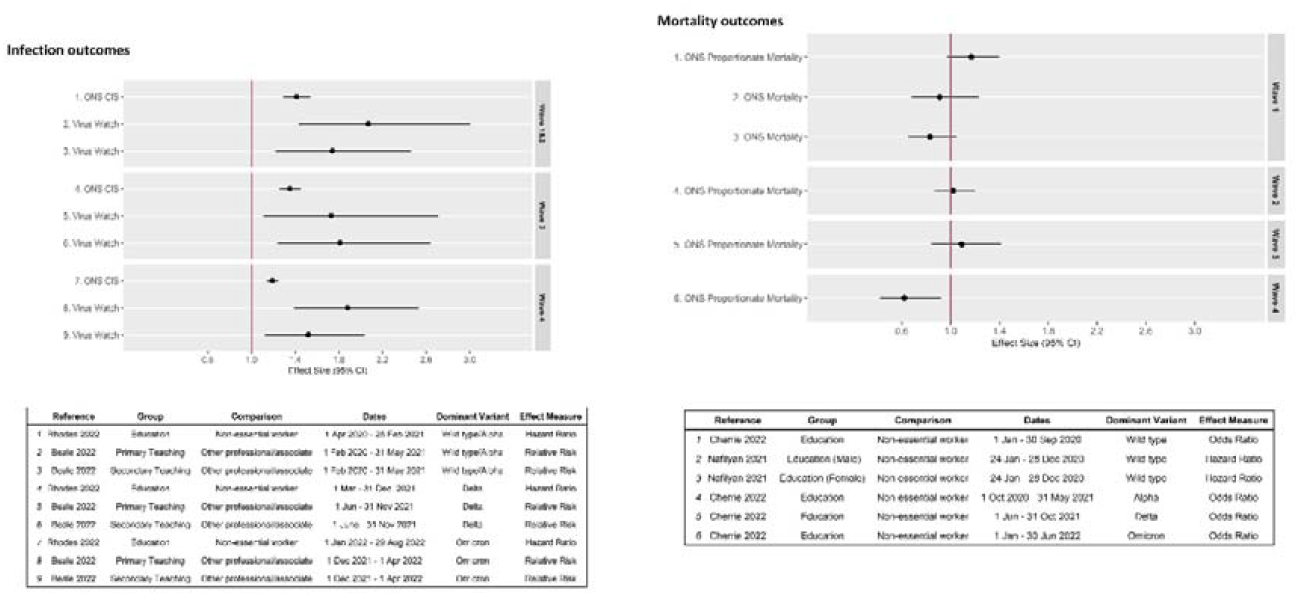
Effect size with 95% confidence interval for infection and mortality outcomes by COVID-19 wave for healthcare workers in the UK.

Figure S5 shows effect sizes and confidence intervals for the transport sector, by wave. There was some evidence of elevated risk during Wave 1 and 2 when combined using the CIS data but not from the Virus Watch data or at other time points. Figure S6 shows relative effects for mortality outcomes by wave for the transport sector. Due to small group sizes the confidence intervals around point estimates for transport were very wide. Results suggested elevated risks of mortality for the transport sector in Wave 1 and Wave 2, but little evidence that this persisted into Waves 3 and 4. Excess mortality analysis saw high levels of excess mortality for transport workers in April 2020.

Table 2 describes the five studies comparing risks and presents key biases and strengths as agreed in the workshop. Three studies were thought to be at risk of bias in the establishment of outcome assessment. Virus Watch largely used the results of self-reported or record-linked test results to establish infection status. The UK testing strategies varied at different points in the pandemic and furthermore, these strategies have been dependent on occupation – for example PCR tests were more widely available for healthcare workers in the early phase of the pandemic while teachers were expected to take regular lateral flow tests between December 2000 and February 2022. In addition, one needs consider that motivation for taking a test may be linked to occupation; a care home worker may be more concerned about attending the workplace while infected than someone who is self-employed and works outdoors. Virus Watch used a serological sub-cohort as a sensitivity analysis that was not prone to the same bias and results were consistent. The ONS CIS study was at lower risk of bias in the ascertainment of outcome as it relied on repeated PCR testing unrelated to occupation. Both the ONS Mortality study and ONS proportionate mortality study used cause of death as reported on death certificates to establish death from COVID-19 and this was believed to put them at risk of bias. Establishing COVID-19 as a cause of death may rely on previous tests (with the risks of bias already mentioned) and/or a subjective judgement from a clinician (9). In addition, there can be uncertainty as to whether a person died from COVID-19 or from other causes while suffering from it. The ONS excess mortality analysis did not utilise cause of death and therefore was not at risk of this type of bias. All three mortality analyses may be missing some outcome data while waiting for coroners reports; particularly amongst healthcare workers.

**Table 2:** Key bias and strengths of epidemiological studies of occupational risks of SARS-CoV-2 infection and COVID-19 mortality funded by the PROTECT National Core Study

| Study short name | Key sources of bias | Strengths |
| --- | --- | --- |
| ONS CIS | Selection bias due to missing study visits – could be linked to long work hours/working away. Likely to attenuate relative risks in healthcare, transport where long hours/shifts likely.<br>Some misclassification of occupation/missing occupation data. Direction of bias unclear but results tested with sensitivity analyses.<br>Confounding due to behaviours outside the workplace. Direction of bias unclear. | Cohort designed to be representative of UK population<br>Use of regular PCR tests independent of occupation.<br>Occupation data regularly updated. |
| Virus Watch | Selection bias due to a self-selecting cohort that is likely to be highly motivated. Protective behaviours that may be related to occupation may attenuate relative differences.<br>Bias in ascertainment of infection via self-reported and linked test results. Likely to elevate relative differences in occupations with more access to testing/requirement for testing such as health, social care and education.<br>Recall bias in reporting of behaviour outside the workplace. May reduce associations.<br>Some misclassification of occupation when occupation changes over time, may attenuate relative risks in transient occupations. | Inclusion of behaviour outside the workplace<br>Use of serology for sensitivity analyses |
| ONS Mortality | Misclassification of occupation in use of 2010 census. May attenuate risk in transient occupations e.g. hospitality.<br>Selection bias in use of 2010 census. May attenuate risks in jobs where migrant workers likely e.g. food production.<br>Bias in ascertainment of cause of death. May lead to exaggerated relative differences in occupations where COVID more likely to be suspected e.g. healthcare.<br>Missing data due to coroners inquests. Likely to attenuate relative differences in | Use of full population data.<br>Adjustment for a series of potential confounders. |
|  | healthcare workers. Known to be small numbers. |  |
| ONS Proportionate mortality | Bias due to lockdown effect on control group. Deaths from other causes likely to reduce in occupations not at work. Likely to attenuate relative risks in occupations working through lockdown such as health, social care.<br>Bias in ascertainment of cause of death. May lead to exaggerated relative differences in occupations where COVID more likely to be suspected e.g. healthcare.<br>Confounding by ethnicity not controlled for in analysis. Proportionate mortality design should take this into account. | Comparison to deaths by other causes likely to minimise confounding due to social factors.<br>Use of death certificate occupation likely to be up to date and include migrant/transient workers. |
| ONS Excess mortality | Lockdown effect likely to reduce mortality from other causes. Likely to attenuate excess mortality.<br>Use of previous 5 year mortality as a predictor of mortality – this naturally fluctuates over time so may not be a good predictor. Unclear how this will vary by occupation. | Not reliant on ascertainment of cause of death<br>Allows examination of time trends in detail. |

Although all observational studies are at risk of unmeasured confounding, two studies were specifically acknowledged to be missing adjustment for key known confounders. The ONS CIS study did not control for behaviour outside the workplace whereas Virus Watch did. The ONS proportionate mortality analysis could not control for ethnic group as this is not recorded on death certificates, whereas the ONS mortality analysis did control for this using the linked census data. The ONS excess mortality made a ‘within-group’ comparison comparing pandemic data to a pre-pandemic average and thus controlling for demographic confounders was not necessary. Note that adjustment for confounding had very little effect on effect estimates in the two studies of infection. In the ONS Mortality analysis, covariate adjustment substantially attenuated hazard ratios; this was not the case for the ONS Proportionate Mortality analysis.

Selection bias was believed to be a concern in three studies. The ONS CIS study has non-response that could be related to occupation e.g. where participants are not available for study visits due to their job. Virus Watch participants are self-selected and their motivations may be linked to occupation. The ONS mortality study utilises 2010 census data and therefore will exclude participants who have entered the UK recently; this could be linked to occupation as migrant workers are likely to be missing.

Bias in the ascertainment of covariates was a cause for concern for both infection studies. The ONS CIS study had some missing occupational data although sensitivity analyses suggested results were robust to this. Virus Watch analyses did not allow for changing occupations with time. This analysis utilised data on behaviour outside the workplace which were felt to be at risk from recall bias. The ONS Mortality study used occupation data from 2010 which was likely to have changed, particularly for the more transient occupations such as hospitality.

The ONS proportionate mortality study and the ONS excess mortality study used ‘within-group’ comparisons which controlled for confounding to some extent, however this may have introduced additional biases. The proportionate mortality compares death from COVID-19 to deaths from other causes; work related deaths from other causes is likely to have been reduced due to lockdown in some cases (e.g. where workers were likely to work from home or be furloughed). The ONS excess mortality compared deaths in 2020/2021 to a five-year pre-pandemic period which may fluctuate by time.

Table 3 shows a set of conclusions we believe can confidently by made as a results of these data, as suggested and agreed by workshop participants and other co-authors.

**Table 3.**
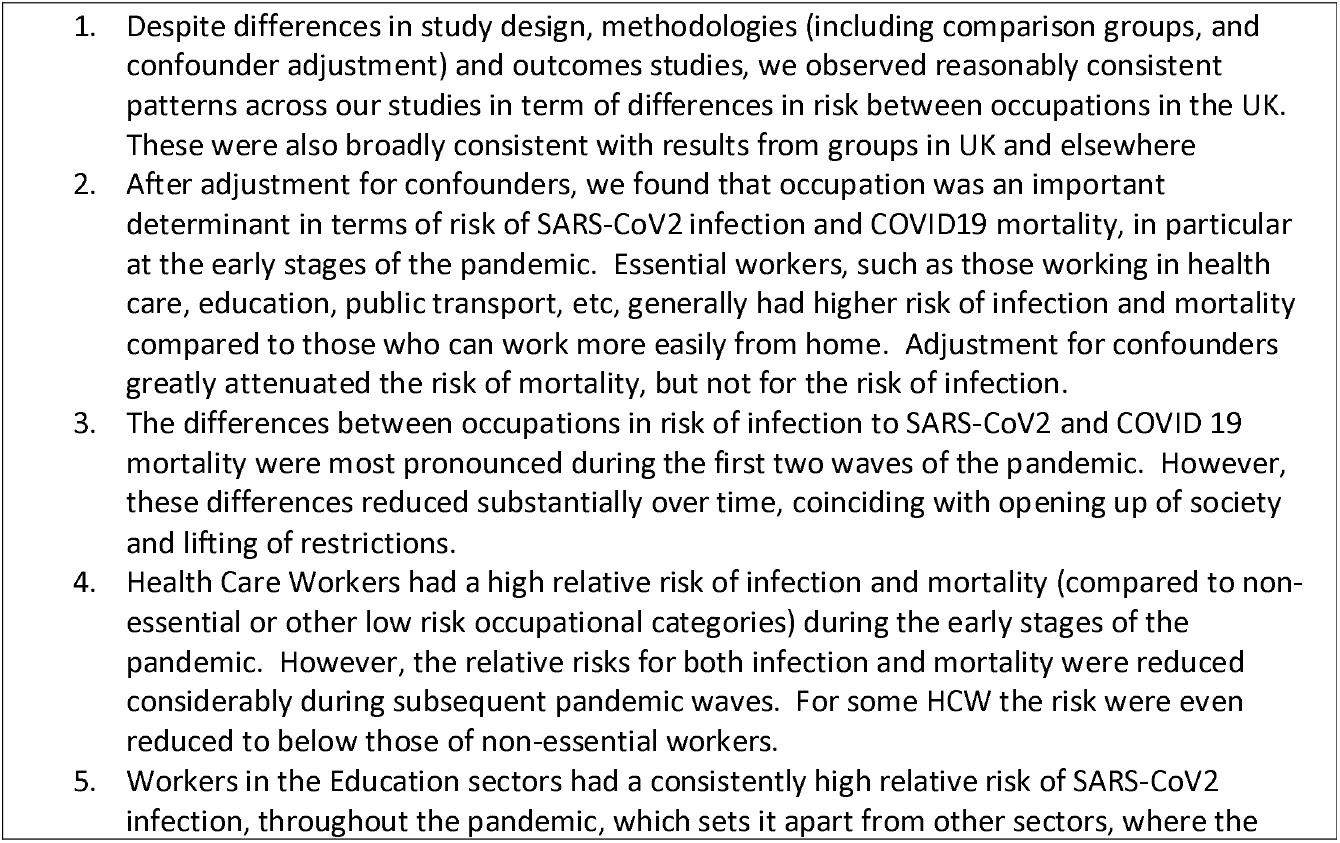
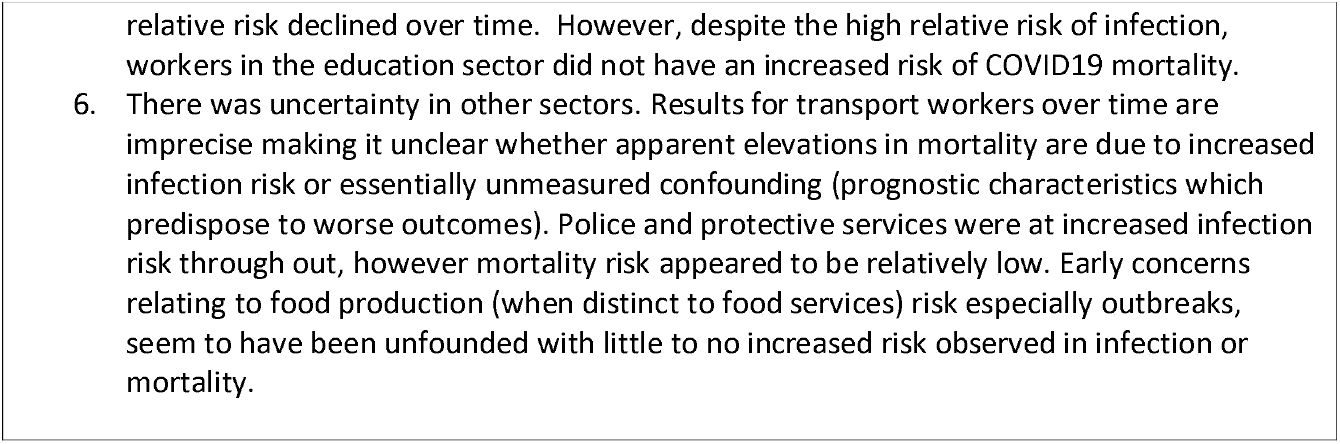
Broad conclusions suggested and agreed by workshop participants

## Discussion

Our results suggest that healthcare workers saw increased risks early in the pandemic but that these were less evident in later waves. We suggest that reasons for this could be a combination of prior infections, enhanced personal protective equipment (PPE), early access to vaccines and high levels of vaccine uptake (10). A survey on workplace mitigation methods conducted as part of the Virus Watch study found that healthcare settings used a wider range of mitigation methods more frequently than other occupational groups, which may have mitigated high risk in these settings; implementation of workplace mitigations was also more persistent in healthcare settings than other workplaces after the relaxation of national restrictions (11). Where there is a delay to these interventions we see a delay in the benefits (lower infection and mortality risk). A good example is the medical support staff who generally had longer periods of poor COVID-19 outcomes compared to more senior staff (3, 5). Therefore, it is important that issues such as vaccine hesitancy, PPE supply or funding are resolved early on and in an equitable way (in terms of occupational equity). A small uptick in relative infection risk was seen in the more contagious Omicron wave; potentially connected to waning immunity and/or relaxed mitigations.

Social care workers saw elevated rates of both infection and mortality, which largely diminished over time. However, while elevated relative effects in the healthcare sector had subsided by Wave 3, elevated risks of mortality appear to have persisted longer for the social care sector. Care workers had some evidence of lower vaccination uptake than other occupations which may have contributed to persistently elevated risk (10). Less intensive and persistent workplace mitigations than in healthcare settings may also have contributed (11).

Education workers were at increased risk of infection in all four waves, but despite this we did not find an elevated risk of mortality. Education workers are known to have high rates of close contact (12) which is likely to explain the sustained infection risk. This mismatch between infection and mortality might suggest unmeasured confounding (i.e. we have not been fully successful in controlling for factors which predispose to severe outcomes, such as morbidity). Other UK researchers (13) found some evidence of increased COVID-19 mortality for secondary school teachers but not for education workers in general. Education workers have high rates of vaccination coverage (10) which may partially temper occupational risk of mortality. We could also speculate that teachers also have strong immune systems from frequent exposure to viral illnesses (14) or a strong bias for healthy workers (15).

There is some evidence of elevated risks of mortality for transport workers but we remain relatively uncertain as to risk of infection for transport workers over time as most results are imprecise. Apparent elevations in mortality may be due to increased infection risk but we cannot rule out unmeasured confounding. Transport workers have been identified in studies of potential risk factors as having relatively low vaccination uptake(16, 17) – including amongst vulnerable workers – as well as lower implementation of some workplace mitigations such as antigen testing programmes(11).

Our results suggest that occupational risk is not static over time in a pandemic for a variety of reasons. The implication is that some occupations go from having low relative risk to higher relative risk between waves. This highlights how important it is to maintain surveillance of cases and mortality, with organisations issuing data and reports to the academic community to analyse periodically. This type of government public healthy capacity, especially if widened to cover other infectious diseases, will be essential for future pandemic preparedness.

Our results were largely consistent across different studies with different biases and strengths. While it would be difficult to predict the direction and magnitude of bias in each study due to multiple biases acting simultaneously, we have used the different study designs to triangulate results (2). When looking at infection risk, the fact that two studies using quite different methodology and with different expected biases yield such similar results is reassuring. When it comes to mortality, we have three studies that address confounding in different ways. The ONS mortality analysis adjust for confounders in a regression model, the proportionate mortality analysis compares death from COVID-19 to deaths from other causes within the same occupation and the ONS excess mortality analysis makes a comparison to a pre-pandemic time period. All three studies reveal similar results for the 2020 period although only two of the studies currently provides results for 2021 onwards. The combination of data from multiple sources and utilising different methodologies has allowed us to increase confidence in our conclusions about high-risk groups and changes over time, and also highlighted some interesting points for further research.

A limitation of this work is that the data extraction, assessment of bias and drawing of conclusions has been carried out largely by the authors of the original studies. We acknowledge that there is subjectivity in all of these processes that may have led to a biased interpretation. An additional limitation is the categorisation of occupation and time-period; these categorisations are a balance between granular detail and statistical power; we are aware that by aggregating data we are likely to have missed differences within categories and time-period. While we see differences between groups we do not know whether these are due to the workplace itself, activities within the workplace or due to unmeasured confounding due to the propensity for certain occupational groups to behave in a particular way outside the workplace.

## Conclusions

There were relative differences between sectors in the UK in occupational COVID-19 risks, which remained after adjustment for confounders. Results were consistent across multiple studies, which increases confidence in conclusions. Largely differences were most pronounced in the early stages of the pandemic and for occupations where workplace attendance was expected. An exception to this was the education sector saw persistent elevation in infection risk with little evidence of increased mortality.

## Data Availability

Data may be obtained from a third party and are not publicly available. The data used in this study are available on the Office for National Statistics (ONS) Secure Research Service for Accredited researchers as the Public Health Research Database. Researchers can apply for accreditation through the Research Accreditation Service.

## Disclaimer

This work was produced using statistical data from ONS. The use of the ONS statistical data in this work does not imply the endorsement of the ONS in relation to the interpretation or analysis of the statistical data. This work uses research datasets which may not exactly reproduce National Statistics aggregates.

From 1 May 2022, Virus Watch received funding from the European Union (Project: 101046314). Views and opinions expressed are however those of the author(s) only and do not necessarily reflect those of the European Union or the European Health and Digital Executive Agency (HaDEA). Neither the European Union nor the granting authority can be held responsible for them.”

## Supplementary materials

**S1.**
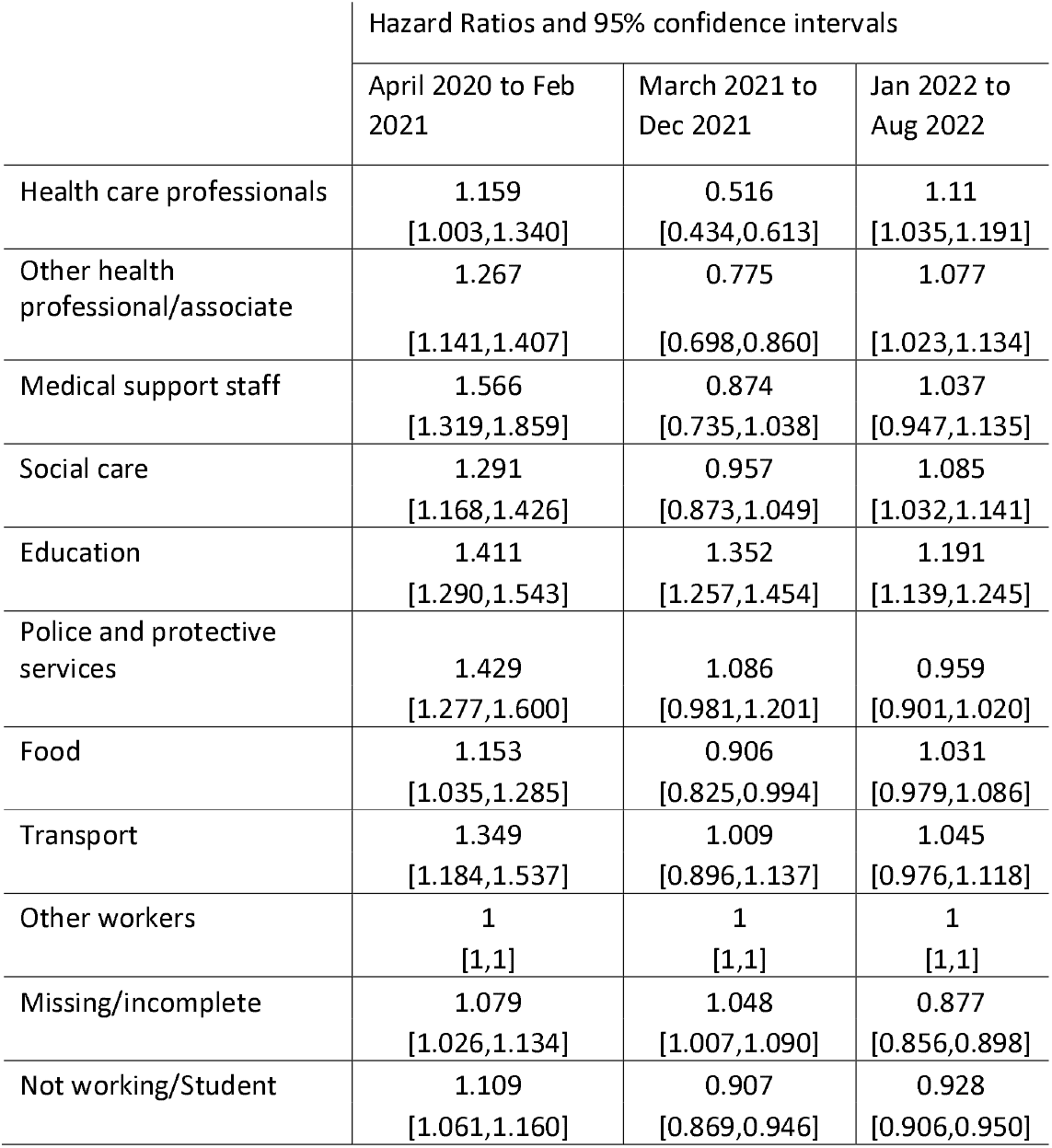
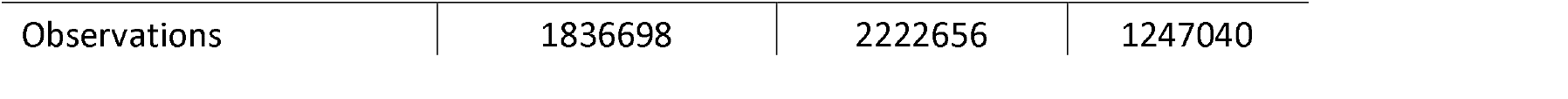
Hazard ratios and 95% confidence intervals from time varying cox regressions for participants aged 20-64 with at least one visit during the time period of interest. Time to positive first positive PCR test as part of CIS survey. All models adjusted for age, sex, ethnicity, IMD, region, rural or urban location, household size and health conditions.

**S2.**
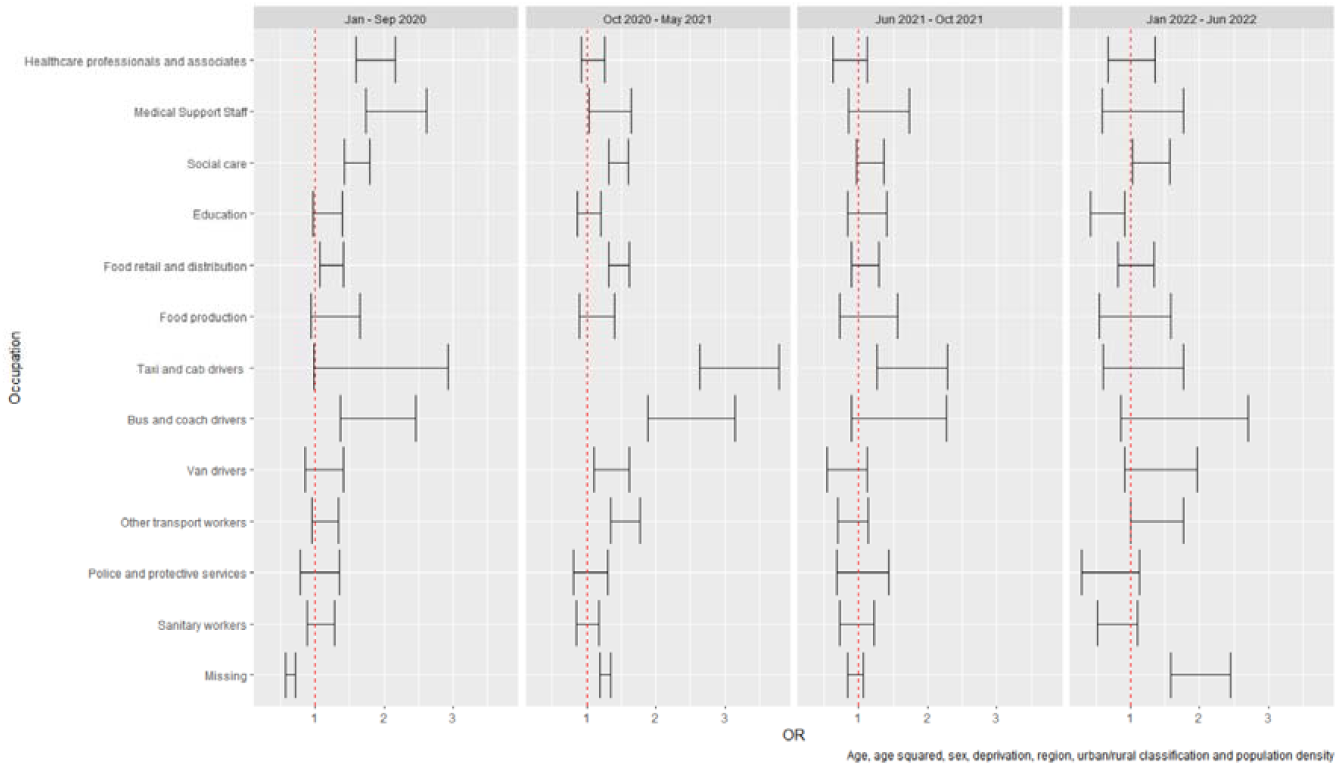
COVID-19 proportionate mortality odds ratios for occupational group (reference – ‘non-essential workers’).

**Figure S3:**
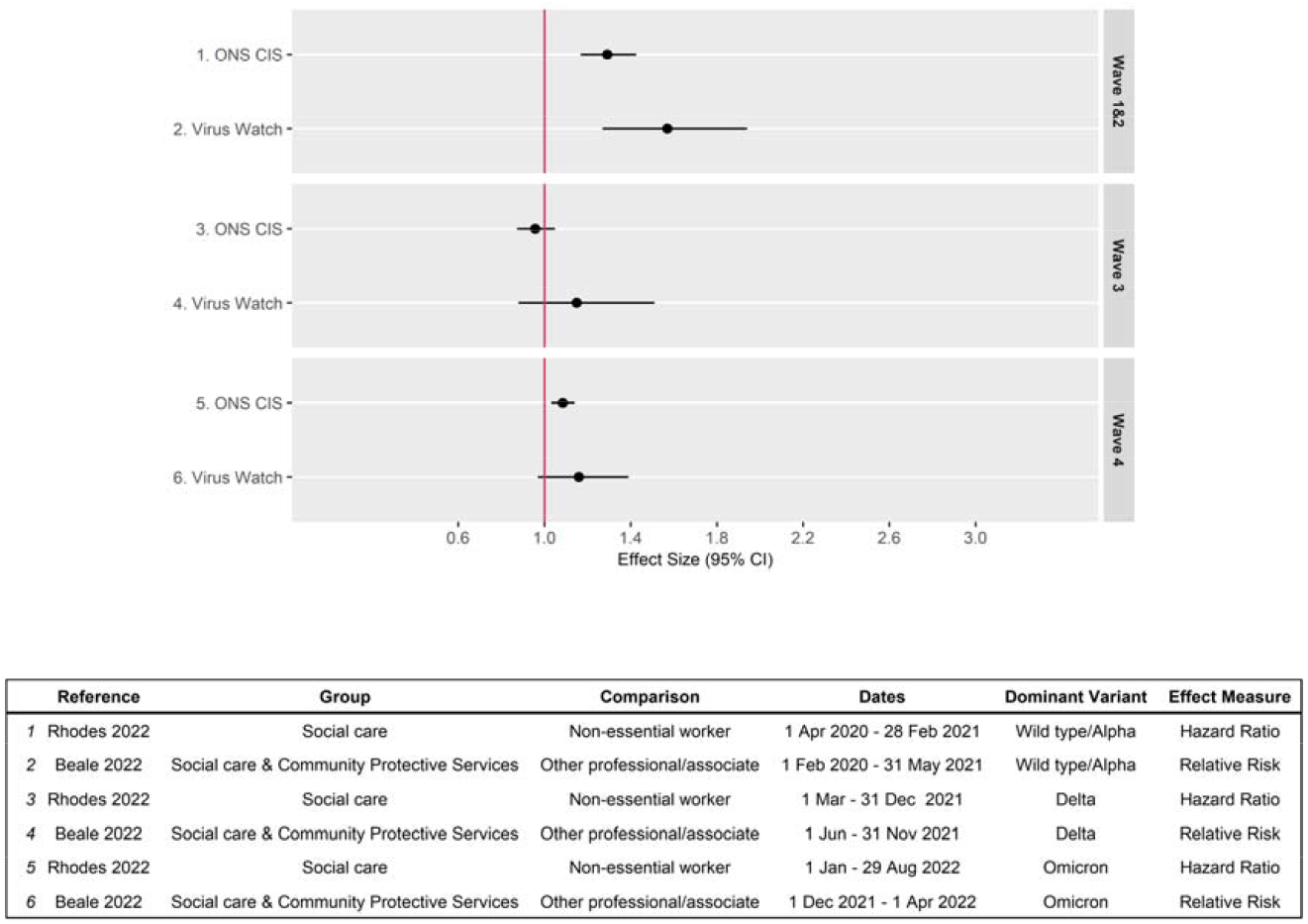
Social care infection by wave

**Figure S4:**
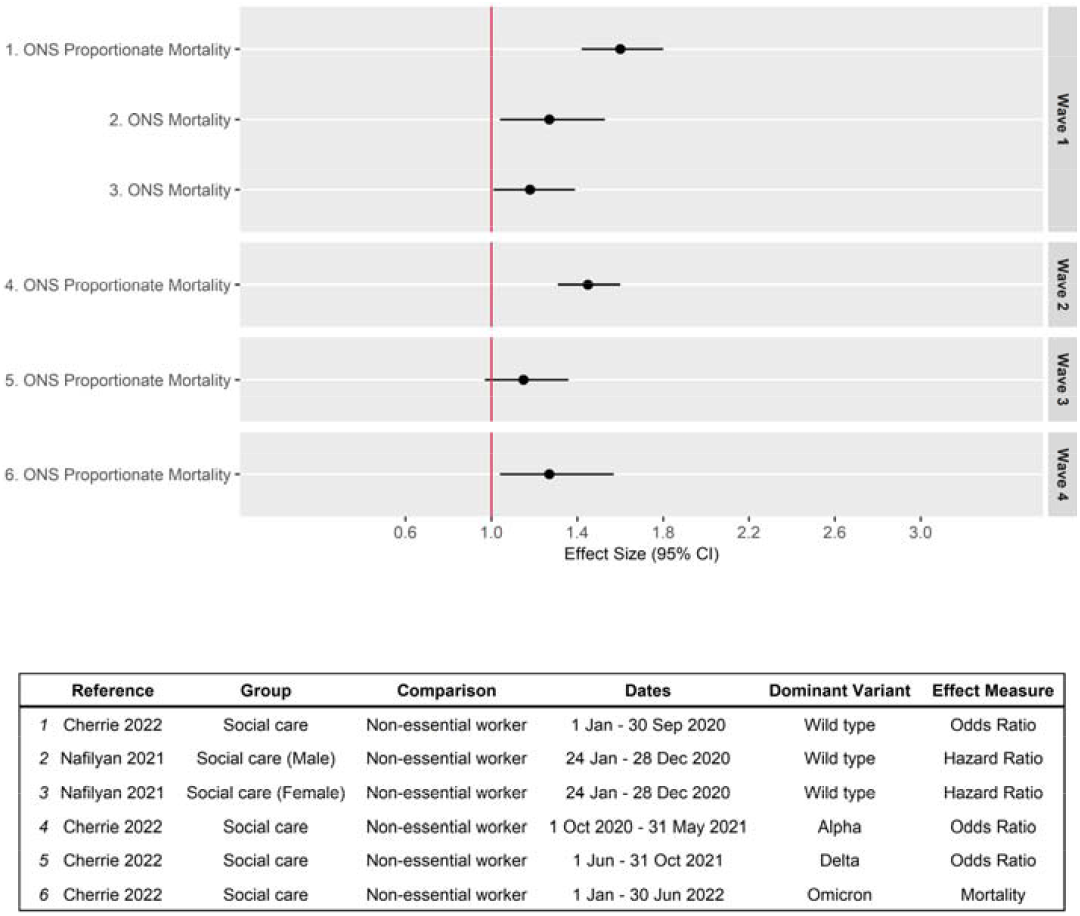
Social care mortality by wave

**Figure S5:**
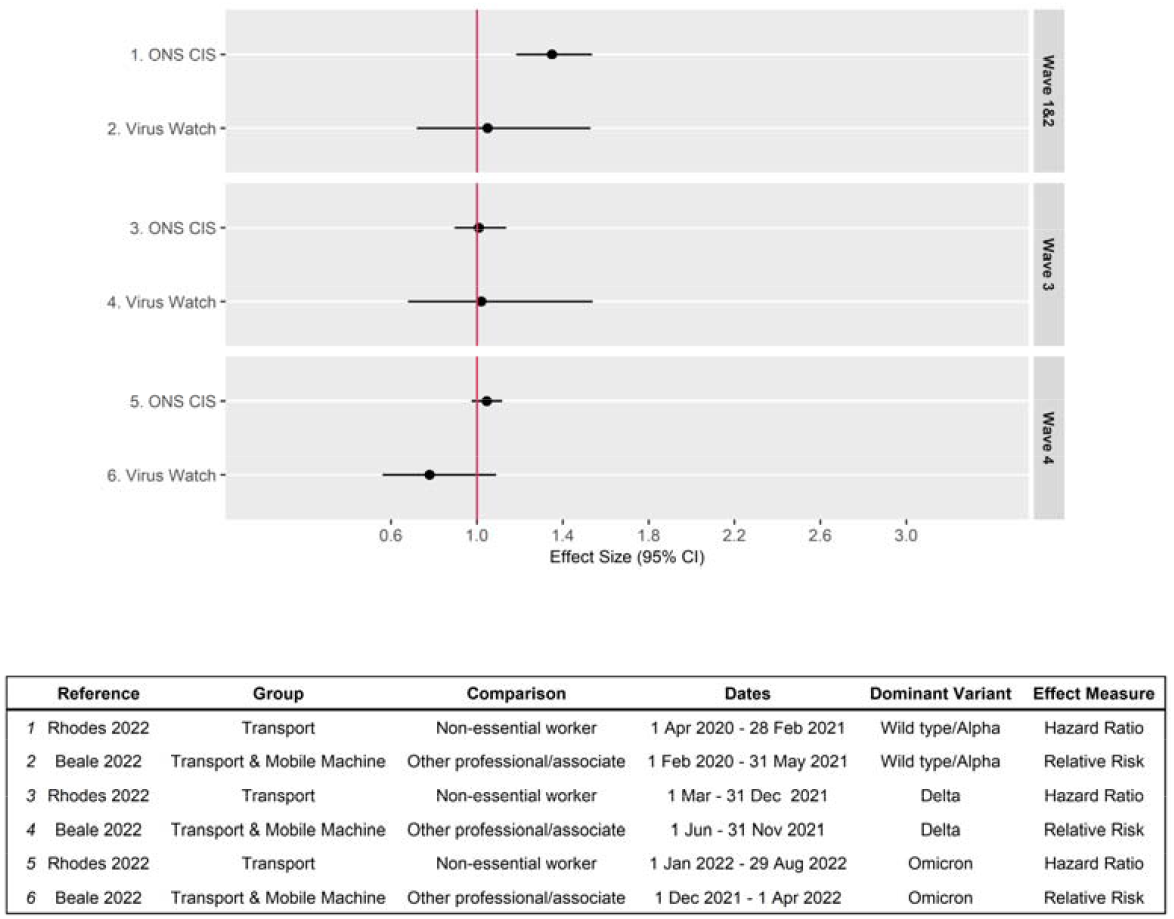
Transport infection

**Figure S6:**
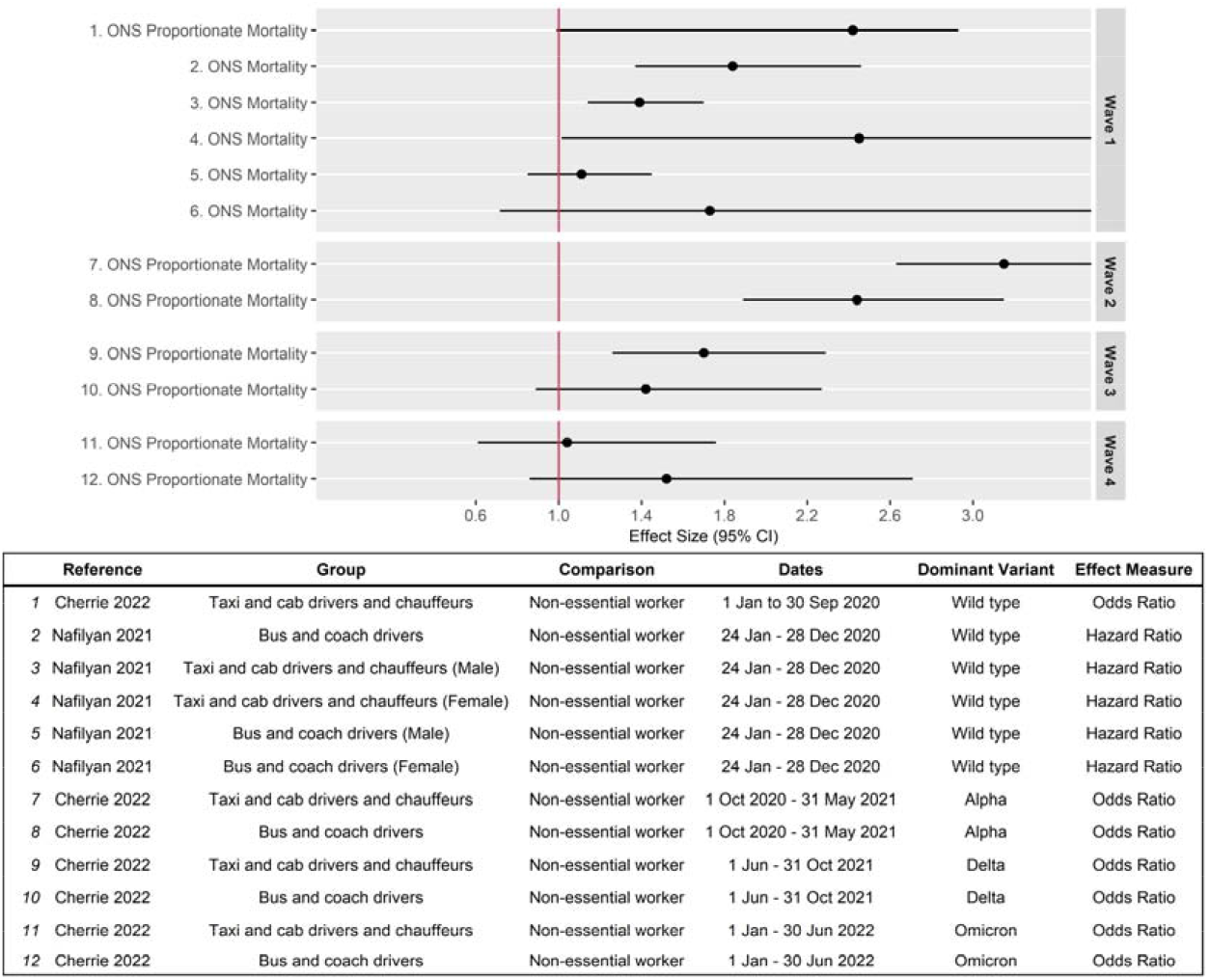
Transport mortality

